# Factors associated with fatal stroke in glioma patients: a population analysis

**DOI:** 10.1101/2021.12.02.21267104

**Authors:** Kai Jin, Paul M Brennan, Michael TC Poon, Cathie LM Sudlow, Jonine D Figueroa

## Abstract

**Importance:** Brain tumour patients have the highest stroke mortality rates among all cancer types, but the factors associated with fatal stroke in brain tumour remain unknown.

**Objective:** We aimed to examine to what extent brain tumour grade, a marker of biological aggressiveness, tumour size and cancer treatment each associated with stroke mortality in glioma. Gliomas include the most common malignant types of brain cancer.

**Design, setting, participants:** A retrospective, observational cohort study using the US National Cancer Institute’s Surveillance Epidemiology and End Results program. We identified adult patients with a primary diagnosis of malignant gliomas in 2000 to 2018 (N=72,252). The primary outcome of interest was death from cerebrovascular disease. Adjusted hazard ratio (aHR) and 95% confidence interval (CI) were calculated using cause-specific Cox regression model to determine associations with tumour characteristics: grades II-IV, tumour size and cancer treatment (surgery, radiotherapy, chemotherapy) associated with stroke mortality after adjustment for age, sex, race, marital status and calendar years.

**Results:** In patients with glioma, increased risk for stroke mortality was observed in patients with higher grade (Grade III: aHR=1.19, 95% CI=0.88-1.61, p>0.05; Grade IV: aHR=1.94, 95% CI=1.39-2.71 compared to Grade II, p<0.001), and those with larger brain tumours (size=3-6 cm: aHR=1.93, 95%CI 1.31 -2.85, p<0.001, size>9cm: aHR=2.07, 95% CI=1.40-3.06, p<0.001 compared to size < 3cm). Having treatment was associated with decreased risk: surgery (yes VS no: aHR= 0.65; p<0.01), radiation (yes VS no: aHR= 0.66, p<0.01), chemotherapy (yes VS no: aHR=0.49, p<0.001).

**Conclusions:** Higher grade and tumour size are strongly associated with increased stroke mortality. This implicates tumour biology and/or the systemic tumour response which require further investigation in prospective studies to determine strategies to mitigate this risk.

## Introduction

Higher stroke mortality rates have been reported for cancer patients compared to non-cancer patients^1^, particularly for brain tumour patients who have the highest risk of fatal stroke among all cancer types^2^. The aetiology of stroke among cancer patients remains unclear and whether stroke is caused by cancer itself or by the treatment of cancer has not been well characterized ^2,3^. Brain tumour-associated stroke may be influenced by tumour biology, direct tumour compression, or cancer therapies^1^. We aimed to examine to what extent brain tumour grades, a marker of biological aggressiveness, tumour size and cancer treatment each associated with fatal stroke in glioma. Gliomas include the most common and most malignant types of brain cancer.

## Methods

We conducted a retrospective, observational cohort study using Surveillance, Epidemiology, and End Results (SEER) according to the Strengthening the Reporting of Observational Studies in Epidemiology (STROBE) guidelines. We identified adult (≥18 years) with primary malignant gliomas between 2000 and 2018 from SEER 18 registries database covering 28% of US population. Glioma is classified into four grades based on WHO criteria^4^. Higher grade indicates increasing tumour aggressiveness. We ascertained cause of deaths by linking to mortality data and the primary outcome of interest was death from cerebrovascular disease using ICD-10 code described previously^3^(see eMethods).

We computed survival time from the date of diagnosis until date of death or last contact (December 31, 2018). Cause-specific cox regression models were used to estimate adjusted hazard ratios (aHRs) and 95% confidence intervals (CIs) to determine factors associated with cerebrovascular mortality among glioma patients including tumour grades (II-IV), tumour size (<=3cm, 3-6 cm, 6-9 cm, >=9 cm), treatment status (surgery yes vs no, radiation therapy yes vs no, chemotherapy yes vs no) adjusted by sociodemographic factors (age, sex,, races/ethnicity, marital status) and calendar years. Due to small numbers of cerebrovascular death in Grade I (N=6) we restricted our analyses to grade II-IV.

Subgroup analyses were performed to check for potential bias and subgroup effects including: 1) age (18-65 years, >65 years); 2) having surgery status; since aging is a strong risk factor for stroke and having surgery is considered a proxy for healthier patients well enough to receive the surgery^5^. We also conducted subgroup analyses by sex (male and female) and ethnicity/race (White, Black). Sensitivity analysis were performed by 1) limiting the study period after 2005 to test the assumption that introduction of adjuvant chemotherapy treatment in 2005 did not influence the probability of occurrence of outcome^6^; 2) limiting one month after diagnosis to reduce the chance of reverse causality^7^. All analyses were performed in R version 4.0. An association was considered statistically significant for a two-sided P value < 0.05.

## Results

Among 72, 252 glioma patients there was a total follow-up time of 266, 491 person-years (median survival=12 months [IQR 4, 34]; 56.8% males) between 2000 to 2018 (Table 1). Fewer patients were diagnosed with low grade (Grade I =2.2% & Grade II =14.6%) and the majority were grade IV, including the most aggressive glioblastoma multiforme (GBM) (61.1%). Most patients were diagnosed younger than 65 years of age, although there was variation by grade (Grade 1: 94.6%; Grade II 83.7%; Grade III 77.4%; Grade IV: 50.4%). Among 374 recorded cerebrovascular deaths, 80% events occurred in Grade III (n=121, 32.3%) and Grade IV gliomas (n=179, 47.9%), and about half in those diagnosed at aged <65 years (Grade III (52.4%) and Grade IV (49.1%).

**Table 1.** Characteristics of the study cohort SEER 2000-2018.

| **Characteristics** | **Total cases** | **Grade I** | **Grade II** | **Grade III** | **Grade IV** | **P value** |
| --- | --- | --- | --- | --- | --- | --- |
| **Number of patients, No.(% of total)** | N=72252 | N=1569 (2.2%) | N=10572 (14.6%) | N=15966 (22.1%) | N=44145 (61.1%) |  |
| **Sex:** |  |  |  |  |  | 0.001 |
| **Female** | 31208 (43.2%) | 736 (46.9%) | 4626 (43.8%) | 7290 (45.7%) | 18556 (42.0%) |  |
| **Male** | 41044 (56.8%) | 833 (53.1%) | 5946 (56.2%) | 8676 (54.3%) | 25589 (58.0%) |  |
| **Age, median (IQR), years** | 59.0 [46.0;70.0] | 31.0 [22.0;44.0] | 46.0 [34.0;59.0] | 50.0 [37.0;64.0] | 64.0 [55.0;73.0] |  |
| **Age categories** |  |  |  |  |  | <0.001 |
| **18-65 years** | 47091 (64.6%) | 1659 (94.6%) | 8936 (83.7%) | 12623 (77.4%) | 23873 (54.0%) |  |
| **66+ years** | 25825 (35.4%) | 95 (5.42%) | 1737 (16.3%) | 3680 (22.6%) | 20313 (46.0%) |  |
| **Year diagnosis** |  |  |  |  |  | 0.001 |
| **2000-2004** | 17640 (24.4%) | 410 (26.1%) | 3112 (29.4%) | 3994 (25.0%) | 10124 (22.9%) |  |
| **2005-2009** | 18715 (25.9%) | 387 (24.7%) | 2984 (28.2%) | 4195 (26.3%) | 11149 (25.3%) |  |
| **2010-2014** | 19843 (27.5%) | 461 (29.4%) | 2575 (24.4%) | 4430 (27.7%) | 12377 (28.0%) |  |
| **2015-2018** | 16054 (22.2%) | 311 (19.8%) | 1901 (18.0%) | 3347 (21.0%) | 10495 (23.8%) |  |
| **Race/ethnicities** |  |  |  |  |  | 0.001 |
| **Non-Hispanic White** | 55079 (76.5%) | 1087 (70.5%) | 7719 (73.3%) | 11485 (72.4%) | 34788 (78.9%) |  |
| **Hispanic (All Races)** | 8553 (11.9%) | 229 (14.9%) | 1497 (14.2%) | 2211 (13.9%) | 4616 (10.5%) |  |
| **Non-Hispanic Black** | 4320 (6.00%) | 136 (8.82%) | 628 (5.97%) | 1065 (6.71%) | 2491 (5.65%) |  |
| **Other ethnic groups** | 4058 (5.64%) | 90 (5.84%) | 683 (6.49%) | 1107 (6.98%) | 2178 (4.94%) |  |
| **Marital status** |  |  |  |  |  | 0.001 |
| **Married/Partner** | 43165 (59.7%) | 619 (39.5%) | 5955 (56.3%) | 8948 (56.0%) | 27643 (62.6%) |  |
| **Single/Separated/Divorced** | 26117 (36.1%) | 866 (55.2%) | 4166 (39.4%) | 6242 (39.1%) | 14843 (33.6%) |  |
| **Unknown** | 2970 (4.11%) | 84 (5.35%) | 451 (4.27%) | 776 (4.86%) | 1659 (3.76%) |  |
| **Tumour size** |  |  |  |  |  | <0.001 |
| **Size <= 3cm** | 7764 (10.6%) | 367 (20.9%) | 1247 (11.7%) | 2196 (13.5%) | 3954 (8.95%) |  |
| **Size 3-6cm** | 20438 (28.0%) | 387 (22.1%) | 2333 (21.9%) | 3415 (20.9%) | 14303 (32.4%) |  |
| **Size 6-9 cm** | 7422 (10.2%) | 68 (3.88%) | 961 (9.00%) | 1412 (8.66%) | 4981 (11.3%) |  |
| **Size more than 9 cm** | 11208 (15.4%) | 297 (16.9%) | 2208 (20.7%) | 3562 (21.8%) | 5141 (11.6%) |  |
| **No data** | 26084 (35.8%) | 635 (36.2%) | 3924 (36.8%) | 5718 (35.1%) | 15807 (35.8%) |  |
| **Surgery performed:** |  |  |  |  |  | 0.001 |
| **Surgery performed** | 52342 (72.4%) | 1377 (87.8%) | 7459 (70.6%) | 10333 (64.7%) | 33173 (75.1%) |  |
| **Surgery not performed** | 19632 (27.2%) | 184 (11.7%) | 3075 (29.1%) | 5554 (34.8%) | 10819 (24.5%) |  |
| **Unknown** | 278 (0.38%) | 8 (0.51%) | 38 (0.36%) | 79 (0.49%) | 153 (0.35%) |  |
| **Radiation** |  |  |  |  |  |  |
| **Radiation given** | 46386 (64.2%) | 238 (15.2%) | 5061 (47.9%) | 9041 (56.6%) | 32046 (72.6%) |  |
| **None/Unknown or Refused** | 25866 (35.8%) | 1331 (84.8%) | 5511 (52.1%) | 6925 (43.4%) | 12099 (27.4%) |  |
| **Chemotherapy** |  |  |  |  |  | <0.001 |
| **Yes** | 36233 (50.1%) | 76 (4.84%) | 3592 (34.0%) | 6535 (40.9%) | 26030 (59.0%) |  |
| **No/Unknown** | 36019 (49.9%) | 1493 (95.2%) | 6980 (66.0%) | 9431 (59.1%) | 18115 (41.0%) |  |
| **Vital status** |  |  |  |  |  | <0.001 |
| **Alive** | 18277 (25.3%) | 1322 (84.3%) | 5210 (49.3%) | 7334 (45.9%) | 4411 (9.99%) |  |
| **Dead** | 53975 (74.7%) | 247 (15.7%) | 5362 (50.7%) | 8632 (54.1%) | 39734 (90.0%) |  |
| **Survival Months** | 12.0 [4.00;34.0] | 87.0 [34.0;149] | 44.0 [12.0;104] | 30.0 [8.00;87.0] | 7.00 [3.00;16.0] | <0.001 |
| **Death from cerebrovascular disease** | 374 | 6 (1.60%) | 68 (18.2%) | 121 (32.4%) | 179 (47.9%) |  |

Increased risk for stroke mortality was observed in patients with high grade gliomas (Grade III: aHR=1.19, 95% CI=0.88-1.61, p>0.05; Grade IV: aHR=1.94, 95% CI=1.39-2.71 compared to Grade II, p<0.001), and those with larger brain tumours (size=3-6 cm: aHR=1.93, 95%CI 1.31 -2.85, p<0.001, size>9cm: aHR=2.07, 95% CI=1.40-3.06, p<0.001 compared to size < 3cm). Having cancer treatment was associated with decreased risk: surgery (yes VS no: aHR= 0.65; p<0.01), radiation (yes VS no: aHR= 0.66, p<0.01), chemotherapy (yes VS no: aHR=0.49, p<0.001) (Figure 1A).

**Figure 1.**
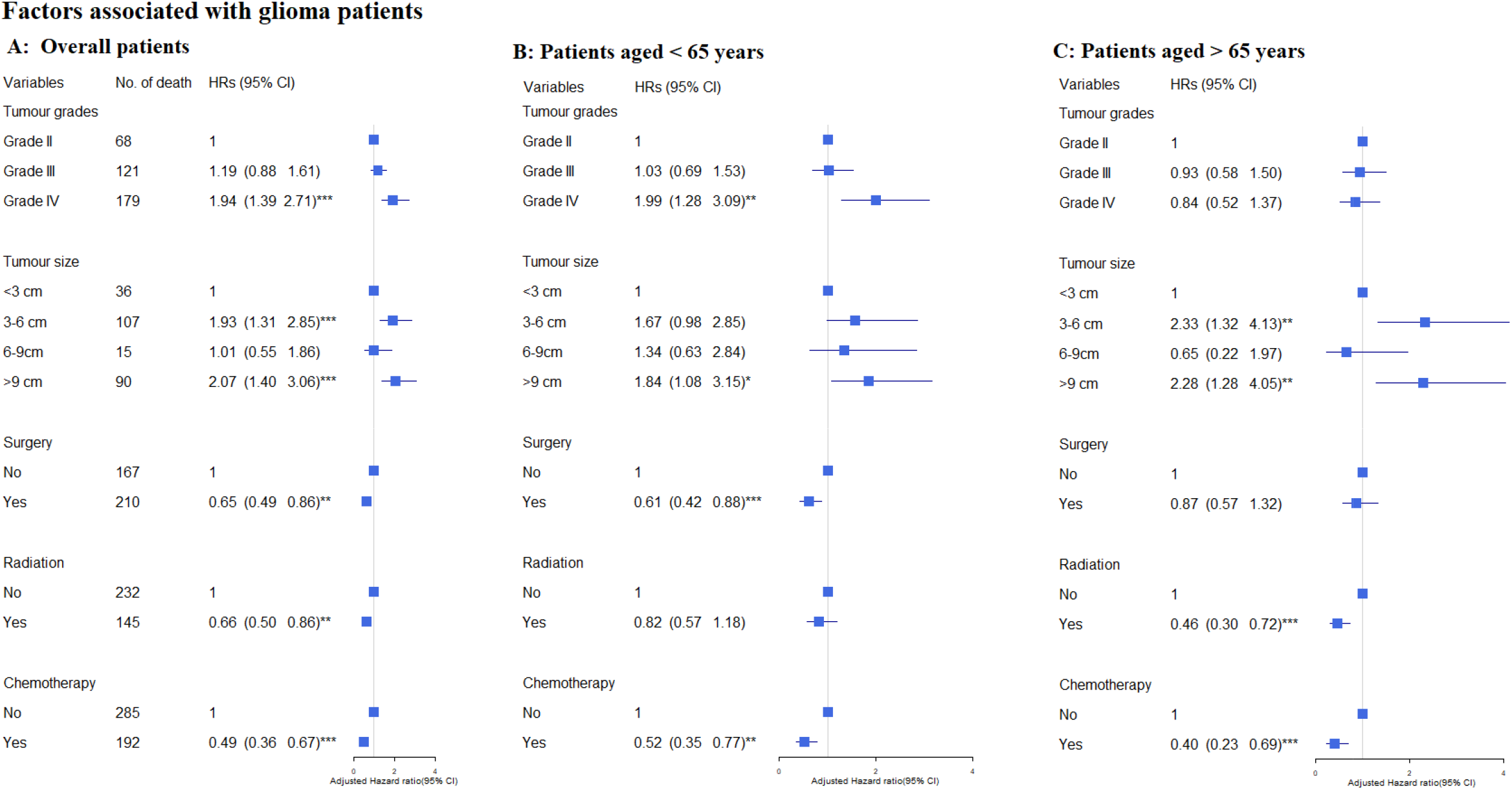
Factors associated with glioma paients. A: overall patients; B: Patients aged <=65 years, C: Patients aged >65 yers. The hazard ratios were calculated using a cause-specific Cox proportional hazards regression model and adjusted by age, sex, calendar year. Significance codes: ‘***’ 0.001, ‘**’ 0.01, ‘*’ 0.05’

In age-stratified analysis, Grade IV (aHR=1.99, 95% CI=1.28-3.09, p<0.01) was strongly associated with stroke morality in patients <65 years (Figure 1B). The aHRs for stroke mortality associated with higher grades appeared to decline in older patients (>65 years) albeit non-significant(Figure 1C). Larger brain tumours were associated with increased stroke mortality, while the more pronounced association was observed in patients aged >65 years. Having cancer treatment was associated lower stroke mortality risk in both age groups. Similar patterns to those noted above were observed in analysis limited to having surgery (eTable1). Broadly comparable estimates were found in sex-stratified analysis and in White although less clear in Black (eTable 2), a study period after 2005 (eTable 3&4) and in those who survived one month after their diagnosis (data not shown).

## Discussion

Our analysis of over 70,000 cases using large population-based data from SEER found higher grade particularly Grade IV, the most aggressive gliomas and larger tumour size were strongly associated with fatal stroke in glioma patients. Receiving cancer treatments was associated with lower risk for fatal stroke.

The association of higher glioma grade with stroke mortality suggests an important biological role for cancer cell aggressiveness in the risk of stroke. This is consistent with previous studies that showed patients with more advanced cancers including lung, pancreatic, colorectal and gastric cancer are also at higher risk of stroke^3,8,9^. This may implicate a systemic response to malignancy in stroke risk, for example from cancer-mediated hypercoagulability status^8,10^. The findings of strong association between high tumour grade and fatal stroke particularly in younger patients may further support the independent role of tumour aggressiveness on the risk of stroke mortality, because younger age group, a relative healthy cohort, was less likely to be affected by comorbidities than older people such as hypertension^11^. The association of larger brain tumours with higher stroke mortality might relate to reduced vascular perfusion from the mass effect of tumour growth, or to the direct tumour invasion into surrounding brain tissue and vasculature, subsequently leading to severe neurological disability or death^12,13^.

Our findings showed that all modalities of cancer treatment were associated with lower risk for fatal stroke. Surgical resection reduces the tumour bulk and helps restore vascular perfusion. Radiotherapy and chemotherapy do not usually impact on the tumour bulk, instead downgrading tumour cell activity. Although cancer treatment attributed to the reduction the risk of fatal stroke in our study, cardiovascular toxic effects of cancer therapies including stroke has received increasing attention. There is evident that cranial radiotherapy was associated with risk of late neurovascular events and stroke in younger brain tumour survivors ^14^. Further study is needed to understand the effects of cancer treatment on subsequent stroke risk to guide preventive intervention^15^.

The complex interplay of putative risk and benefit from the tumour and its treatment underscore the further analysis which is needed. Our findings should be interpreted in the context of the limitations. A lack of clinical variables such as cardiovascular risk factors, cancer treatment regimen, and stroke subtypes as ischaemic or haemorrhagic in the database examined limits further interpretation. A more comprehensive approach is needed by linking health data from various sources.

## Data Availability

The data are available and can be accessed through the SEER database, which is publicly available at https://seer.cancer.gov/data/

## Figure legend

**Figure 1** Cause-specific Cox proportional hazard regression model HRs for association between tumour grades, tumour size, cancer treatment and stroke mortality in gliomas patients and by age groups

