## Supplemental file for "Factors associated with fatal stroke in glioma patients: a population analysis"

**eMethods:** expanded methods

**eTable 1.** Cause-specific Cox proportional hazard regression model HRs for association between tumour grades, tumour size, cancer treatment and stroke mortality in gliomas patients having surgery and by age group between 2000-2008

**eTable 2**. Cause-specific Cox proportional hazard regression model HRs for association between tumour grades, tumour size, cancer treatment and stroke mortality in gliomas patients stratified by sex and ethnicity/race between 2000-2008

**eTable 3.** Cause-specific Cox proportional hazard regression model HRs for association between tumour grades, tumour size, cancer treatment and stroke mortality in gliomas patients and by age group after 2015

**eTable 4.** Cause-specific Cox proportional hazard regression model HRs for association between tumour grades, tumour size, cancer treatment and stroke mortality in gliomas patients having surgery and by age group after 2015

### eMethods

#### Study design

We conducted a retrospective, observational cohort study using Surveillance, Epidemiology, and End Results (SEER) according to the Strengthening the Reporting of Observational Studies in Epidemiology (STROBE) guidelines. We examined the association of brain tumour grades, tumour size and cancer treatment with stroke mortality in patients with glioma using SEER.

#### Data sources

SEER have been previously described[1]. SEER is a registry of population-based incident tumor registries from geographically distinct regions in the USA, covering 28% of the US population, including incidence, survival, and surgical treatment data. The SEER registry does not code comorbidities, performance status, surgical pathology, margin status, doses, or agents. The SEER database is representative of the population of the USA, and this has been validated by external studies[2-5]. Mortality data are coded using ICD classification scheme and are derived from death certificate data. There could be some misclassification due to the coding of cause of death due to the index-cancer may not be perfect6.

**Study population and grading of gliomas**

We identified adult patients (≥18 years) with primary malignant gliomas between 2005 and 2018 from SEER 18 registries database. We classified patients with glioma using the following International Classification of Diseases 10th revision (ICD-10) codes: C700-C729. We used the International Classification of Diseases for Oncology third edition (ICD-O-3) codes to group glioma following the definitions from the Central Brain Tumour Registry of the United States (CBTRUS)descripted prescously[7]. Gliomas is classified based on histological and molecular type[8]. Based on WHO criteria, glioma is classified into four grades and higher grade indicates increasing tumour aggressiveness[9]: Grade I incudes pilocytic astrocytoma , Grade II includes low grade diffuse astrocytoma, Grade III includes anaplastic astrocytoma and Grade IV includes the most aggressive and malignant glioblastoma multiforme (GBM). Histology codes were following the definitions from the Central Brain Tumour Registry of the United States (CBTRUS) described previously[1]. Although pilocytic astrocytoma (Grade I) classified as a non-malignant tumour by WHO, this histology has been historically classified as malignant for mandatory US cancer registry reporting [10].

**Statistical analysis**

Descriptive analyses of baseline characteristics in glioma and by grades were performed using the Pearson $\chi 2$ test for categorical variables (summarized as frequencies/percentages). Continuous variables were compared across subgroups using analysis of variance for normal distribution data presented as mean, 95% confidence interval (CI), or Kruskal–Wallis (summarized as medians and interquartile range) for non-normal distribution.

The primary outcome of interest was death from cerebrovascular disease using ICD 10 code described previously [1], exposures of interest were tumour grades, tumour size and treatment status. Survival analysis was performed and assessed by Kaplan-Meier methods and compared by the log-rank test to access cerebrovascular-cause specific mortality. Deaths from other causes were censored at the time of death. We computed survival time from the date of diagnosis until date of death or last contact (December 31, 2018) and proportional hazards assumption was tested with the Schoenfeld residuals. Due to small numbers of cerebrovascular death in Grade 1 (N=6) we restricted analysis to grades II-IV in the final analysis. Cause-specific cox regression models were used to estimate adjusted hazard ratios (HRs) and 95% confidence intervals (CIs) to determine factors associated with cerebrovascular mortality among glioma patients including tumour grades, tumour size (<=3cm, 3-6 cm, 6-9 cm, >=9 cm), treatment status (surgery yes vs no , radiation therapy yes vs no, chemotherapy yes vs no), adjusted by sociodemographic factors (age , sex, , races/ethnicity, marital status) and calendar years.

Subgroup analyses were performed to check for potential bias and subgroup effects including: 1) age (18- 65 years, >65 years); 2) having surgery. This was to test the assumption that cerebrovascular mortality were not affected by older age that often accompanied by comorbidities and/or risk for stroke [6] while younger age was usually healthy with less comorbidities[7]; having surgery may be a proxy for healthier patients generally well enough to receive the surgery[8], thus representing a relative healthy cohort.[11]. We also conducted subgroup analyses by sex (male and female) and ethnicity/race (White, Black, not performed in Hispanic and other ethnics due to small number of cases). Sensitivity analysis were performed by 1) limiting the study period after 2005 to test the assumption that introduction of adjuvant chemotherapy treatment in 2005 did not influence the probability of occurrence of outcome[12]; 2)for age group of 18-60 years and >60 years old; 3) landmark analyses to those with follow-up commencing 1 month after cancer diagnoses to reduce the chance of reverse causality thereby excluding patients with an event (death or cerebrovascular disease event) within 1 months of cancer diagnosis[13]. All analyses were performed in R version 4.0. An association was considered statistically significant for a two-sided P value < 0.05.

**eTable 1. Cause-specific Cox proportional hazard regression model HRs for association between tumour grades, tumour size, cancer treatment and stroke mortality in gliomas patients having surgery**

| Characteristics | Overall having surgery | By age group | |
|---|---|---|---|
| | | Age<65 with surgery | Age> 65 with surgery |
| | Adjusted HR[a] (95% CI) | Adjusted HR[a] (95% CI) | Adjusted HR[a] (95% CI) |
| **Grades** | | | |
| Grade 2 | **Reference** | **Reference** | **Reference** |
| Grade3 | 1.30 (0.86 1.96) | 1.04 (0.63 1.71) | 1.05 (0.50 2.19) |
| Grade 4 | 2.87 (1.80 4.56)*** | 2.27 (1.29 4.00)** | 1.05 (0.50 2.21) |
| **Tumour size** | | | |
| <=3 cm | **Reference** | **Reference** | **Reference** |
| 3-6 cm | 2.04 (1.17 3.54)* | 2.09 (0.96 4.54) | 2.42 (1.08 5.42)* |
| 6-9cm | 1.22 (0.55 2.70) | 2.11 (0.81 5.49) | 0.36 (0.04 2.87) |
| >9 cm | 2.29 (1.28 4.10)*** | 2.94 (1.34 6.46)** | 1.65 (0.68 3.99) |
| **Receiving radiation** | | | |
| No | **Reference** | **Reference** | **Reference** |
| Yes | 0.81 ( 0.58 1.14) | 1.05 (0.67 1.64) | 0.55 (0.32 0.95)* |
| **Receiving chemotherapy** | | | |
| No | **Reference** | **Reference** | **Reference** |
| Yes | 0.36 ( 0.24 0.52)*** | 0.43 (0.27 0.70)*** | 0.25 (0.13 0.50)*** |

The hazard ratios were calculated using a cause-specific Cox proportional hazards regression model and adjusted by age, sex, ethnicity/race, marital status, calendar year. Signif. codes: '***' 0.001, '**' 0.01, '*' 0.05 '

**eTable 2 Cause-specific Cox proportional hazard regression model HRs for association between tumour grades, tumour size, cancer treatment and stroke mortality in gliomas patients stratified by sex and ethnicity/race**

| Characteristics | By sex | | By race | |
|---|---|---|---|---|
| | Female | Male | White | Black |
| | Adjusted HR[a] (95% CI) | Adjusted HR[a] (95% CI) | Adjusted HR[a] (95% CI) | Adjusted HR[a] (95% CI) |
| **Grades** | | | | |
| Grade 2 | **Reference** | **Reference** | **Reference** | **Reference** |
| Grade3 | 1.27 (0.82 1.97) | 1.09 (0.72 1.67) | 1.25 (0.86 1.80) | 0.72 ( 0.31 1.70) |
| Grade 4 | 1.40 (0.84 2.33) | 2.45 (1.57 3.82)*** | 2.09 (1.39 3.13)*** | 1.59 (0.63 4.01) |
| **Tumour size** | | | | |
| <=3 cm | **Reference** | **Reference** | **Reference** | **Reference** |
| 3-6 cm | 2.05 ( 1.15 3.67)* | 1.81 (1.07 3.05)* | 1.57 (1.01 2.45)* | 1.94 (0.59 6.32) |
| 6-9cm | 0.73 (0.24 2.21) | 1.12 (0.53 2.37) | 0.57 (0.25 1.30) | NA |

| | | | | |
|---|---|---|---|---|
| >9 cm | 2.25 (1.26 4.01)** | 1.90 (1.12 3.24)* | 1.84 (1.19 2.85)** | 2.71 (0.88 8.41) |
| **Receiving surgery** | | | | |
| No | **Reference** | **Reference** | **Reference** | **Reference** |
| Yes | 0.65 (0.43 0.98)* | 0.66 (0.46 0.95)* | 0.69 (0.50 0.96)* | 0.48 (0.22 1.05) |
| **Receiving radiation** | | | | |
| No | **Reference** | **Reference** | **Reference** | **Reference** |
| Yes | 0.79 (0.53 1.18) | 0.57 (0.39 0.82)** | 0.58 (0.42 0.81)** | 0.57 (0.25 1.30) |
| **Receiving chemotherapy** | | | | |
| No | **Reference** | **Reference** | **Reference** | **Reference** |
| Yes | 0.55 (0.35 0.88)* | 0.45 (0.30 0.68)*** | 0.48 (0.33 0.70)*** | 0.58 (0.21 1.60) |

The hazard ratios were calculated using a cause-specific Cox proportional hazards regression model and adjusted by age, sex, ethnicity/race, marital status, calendar year. Significance codes: '***' 0.001, '**' 0.01, '*' 0.05 '

**eTable 3 Cause-specific Cox proportional hazard regression model HRs for association between tumour grades, tumour size, cancer treatment and stroke mortality in gliomas patients and by age group after 2005**

| **Characteristics** | **Overall** | **<65 years** | **>65 years** |
|---|---|---|---|
| | **Adjusted HR[a]** | **Adjusted HR[a]** | **Adjusted HR[a]** |
| | **(95% CI)** | **(95% CI)** | **(95% CI)** |
| **Grades** | | | |
| **Grade 2** | **Reference** | **Reference** | **Reference** |
| **Grade3** | 1.15 (0.78 1.70) | 1.15 (0.67 1.97) | 0.79 (0.45 1.38) |
| **Grade 4** | 1.72 (1.12 2.63)* | 2.33 (1.30 4.17)** | 0.64 (0.36 1.16) |
| **Tumour size** | | | |
| **<=3 cm** | **Reference** | **Reference** | **Reference** |
| **3-6 cm** | 1.95 (1.30 2.92)** | 1.58 (0.90 2.77) | 2.51 (1.38 4.57)** |
| **6-9cm** | 1.00 ( 0.53 1.89) | 1.27 (0.58 2.79) | 0.72 (0.23 2.20) |
| **>9 cm** | 2.05 (1.37 3.08)*** | 1.75 (1.00 3.06)* | 2.40 (1.33 4.35)** |
| **Receiving surgery** | | | |
| **No** | **Reference** | **Reference** | **Reference** |
| **Yes** | 0.58 (0.41 0.82)** | 0.48 (0.30 0.76)** | 0.88 (0.51 1.52) |
| **Receiving radiation** | | | |
| **No** | **Reference** | **Reference** | **Reference** |
| **Yes** | 0.66 (0.46 0.96)* | 0.73 (0.44 1.21) | 0.55 (0.31 0.98)* |
| **Receiving chemotherapy** | | | |
| **No** | **Reference** | **Reference** | **Reference** |
| **Yes** | 0.49 (0.33 0.74)*** | 0.49 (0.29 0.85)* | 0.41 (0.21 0.79)** |

**eTable 4. Cause-specific Cox proportional hazard regression model HRs for association between tumour grades, tumour size, cancer treatment and stroke mortality in gliomas patients having surgery and by age group after 2005**

| Characteristics | Overall having surgery | By age groups | |
|---|---|---|---|
| | | Age<65 with surgery | Age> 65 with surgery |
| | Adjusted HR[a] | Adjusted HR[a] | Adjusted HR[a] |
| | (95% CI) | (95% CI) | (95% CI) |
| **Grades** | | | |
| **Grade 2** | **Reference** | **Reference** | **Reference** |
| **Grade3** | 1.01 (0.59 1.72) | 0.73 (0.36 1.46) | 0.91 (0.38 2.19) |
| **Grade 4** | 2.25 (1.25 4.06)** | 2.32 (1.11 4.86)* | 0.72 (0.29 1.79) |
| **Tumour size** | | | |
| **<=3 cm** | **Reference** | **Reference** | **Reference** |
| **3-6 cm** | 1.82 ( 1.03 3.19)* | 1.68 (0.76 3.71) | 2.37 (1.04 5.38)* |
| **6-9cm** | 1.07 (0.47 2.44) | 1.77 (0.66 4.76) | 0.35 (0.04 2.83) |
| **>9 cm** | 2.11 (1.17 3.80)* | 2.71 (1.22 6.03)* | 1.35 (0.54 3.35) |
| **Receiving radiation** | | | |
| **No** | **Reference** | **Reference** | **Reference** |
| **Yes** | 0.82 (0.51 1.31) | 0.78 (0.41 1.48) | 0.77 (0.39 1.54) |
| **Receiving chemotherapy** | | | |
| **No** | **Reference** | **Reference** | **Reference** |
| **Yes** | 0.33 (0.20 0.54)*** | 0.42 (0.21 0.81)** | 0.20 (0.09 0.45)*** |

The hazard ratios were calculated using a cause-specific Cox proportional hazards regression model and adjusted by age, sex, ethnicity/race, marital status, calendar year. Signif. codes: '***' 0.001, '**' 0.01, '*' 0.05 '
